# Integrating Plasma Brain-Derived pTau217, White Matter Hyperintensity and Hippocampal Volumes as Optimized Prognostic Markers for Cognitive Decline in an Asian Cohort with High Cerebrovascular Disease Burden

**DOI:** 10.64898/2026.11.23.26368742

**Authors:** Joyce R. Chong, Saima Hilal, Thomas K. Karikari, Narayanaswamy Venketasubramanian, Michael Schöll, Henrik Zetterberg, Sterling C. Johnson, Christopher P. Chen, Mitchell K.P. Lai

## Abstract

**Background:** Combination of plasma and neuroimaging biomarkers associated with Alzheimer’s disease (AD) and cerebrovascular pathophysiology might provide more accurate prediction of cognitive decline in populations with extensive AD and cerebral small vessel disease copathologies. Here, we aimed to identify an optimized biomarker panel for predicting cognitive decline by evaluating combinations of plasma and magnetic resonance imaging (MRI) biomarkers.

**Methods:** We investigated participants without dementia or with mild dementia, recruited from memory clinics and the community in Singapore between August 2010 and August 2020. Plasma biomarkers of brain amyloid (brain-derived [BD]-pTau217, BD-pTau181, BD-pTau231, total-pTau217, total-pTau181, and total-pTau231), astrogliosis (GFAP), and neuroaxonal injury (NfL), with MRI measures of cerebrovascular disease (white matter hyperintensity [WMH] volume) and neurodegeneration (hippocampal volume), and primary outcome of longitudinal Clinical Dementia Rating Sum of Boxes (CDR-SB; mean [SD] follow-up duration, 51.0 [15.8] months). All plasma and MRI biomarker combinations for predicting CDR-SB slope (annual change in CDR-SB) were evaluated.

**Results:** This study included 523 participants (mean [SD] age, 72.4 [7.9] years; 291 females [55.6%]). The parsimonious model included plasma BD-pTau217, WMH volume, and hippocampal volume (all *P* ≤ 0.004). Dichotomizing these biomarkers yielded A, V, and N status, respectively, classifying participants as all negative (A-V-N-; n = 128), 1-positive (A+V-N-, A-V+N- or A-V-N+; n = 154), 2-positive (A+V+N-, A+V-N+, A-V+N+;n = 158) or 3-positive (A+V+N+; n = 83). Annual CDR-SB progression increased stepwise with the number of positive biomarkers. Compared with the all negative group (β, 0.016), the 1-positive (β, 0.409), 2-positive (β, 1.16) and 3-positive groups (β, 1.49) showed faster cognitive decline (all *P* < 0.001). Moreover, the 2-positive and 3-positive groups progressed faster than the 1-positive group (both *P* < 0.001), and the 3-positive group declined faster than 2-positive group (*P* = 0.009).

**Conclusions:** Presence of multiple pathophysiological processes was associated with greater cognitive decline, highlighting the utility of a comprehensive multimodal biomarker profile to enhance prognostication.

## Background

Recent technological advances have enabled the rapid development of blood-based biomarkers associated with Alzheimer’s disease (AD) pathology. In particular, plasma phosphorylated tau217 (pTau217) has emerged as the most promising biomarker for AD pathology. It has been widely validated across diverse cohorts, demonstrating high accuracy in identifying individuals with abnormal brain amyloid burden [1] and predicting subsequent cognitive decline [2–4]. More recent studies have shown that plasma brain-derived pTau217 (BD-pTau217), which more specifically measures pTau originating in the brain, offers superior diagnostic performance compared with conventional total-pTau217, which measures pTau from both the brain and the periphery [5, 6]. Although plasma BD-pTau217 has also demonstrated prognostic utility for cognitive decline [5], a head-to-head comparison of its prognostic performance with that of total-pTau217 remains lacking.

While plasma pTau217 provides prognostic information related to AD, previous autopsy [7–9] and clinical studies [10–12] have reported varying prevalence of concomitant non-AD pathologies, such as cerebrovascular disease (CeVD), that may also contribute to cognitive decline [8, 12, 13]. Accordingly, the recent revised criteria from the Alzheimer’s Association Workgroup recommended a multimodal biomarker profile that reflects an individual’s neuropathophysiological state beyond the presence of AD [14]. For example, such profiles could incorporate biomarkers of AD pathology, including amyloid (“A” or ”T1”) and tau (“T2”), as well as biomarkers of neurodegeneration (“N”), inflammation (“I”), CeVD (“V”), and/or α-synuclein (“S”). Clinically, comprehensive biomarker profiles could help determine the etiology of symptoms and inform treatment decisions. Importantly, they could also improve the prediction of cognitive decline and identify individuals at increased risk of clinical progression in the near term.

Most previous clinical studies examining the effects of AD and co-pathologies on cognitive decline were conducted in White populations [8, 12, 13, 15–18]. AD pathology was assessed using positron emission tomography (PET), cerebrospinal fluid (CSF) tests, and/or plasma pTau217. Non-AD pathologies include CeVD, assessed using magnetic resonance imaging (MRI) measures such as white matter hyperintensity (WMH) volume [13, 16, 18], and α-synucleinopathy, assessed using CSF α-synuclein seed amplification assays [12, 15]. Studies have also examined other key pathophysiological processes implicated in neurodegenerative dementias, including inflammation, assessed using biomarkers such as plasma glial fibrillary acidic protein (GFAP) [17, 19], and neurodegeneration, assessed using biomarkers such as MRI-derived hippocampal volume [18] and plasma neurofilament light chain (NfL) [17]. However, a comprehensive evaluation of the optimal multimodal biomarker combination for predicting cognitive decline and clinical progression in Asian cohorts remains lacking, particularly in cohorts characterized by high prevalence of CeVD. Leveraging plasma biomarkers (BD-pTau and total-pTau species, GFAP, and NfL), MRI biomarkers (WMH volume and hippocampal volume), and longitudinal cognitive data from a large Singapore-based cohort, which has previously been reported to have a high prevalence of CeVD, we aimed to determine the optimal biomarker combination for predicting cognitive decline and clinical progression. We hypothesized that the optimal prognostic panel would include biomarkers reflecting the cohort’s key neuropathological features, namely amyloid (“A”) and CeVD (“V”), as well as non-specific but mechanistically important processes that drive cognitive decline, such as neurodegeneration (“N”).

## Methods

### Study Participants

This study included participants from the Harmonisation cohort, which recruited 700 older adults between August 2010 and August 2020 from memory clinics at the National University Hospital and St Luke’s Hospital in Singapore, as well as from the community. Descriptions of the cohort have been previously published [20, 21]. Briefly, participants underwent annual follow-up for up to 5 years, including serial cognitive assessments with the Clinical Dementia Rating (CDR). At baseline, demographic information and clinical risk factors were determined through a detailed questionnaire, clinical interviews, and medical record searches. Apolipoprotein E ε4 (APOE ε4) carrier status was defined as having at least one ε4 allele. Participants also completed the Mini-Mental State Examination (MMSE). The present study included only participants without dementia or with mild dementia, defined by a CDR Global Score (CDR-GS) of 0, 0.5, or 1. The study cohort flow chart is shown in Supplemental Figure 1.

### Longitudinal Cognitive Trajectories and Incident Cognitive Decline

Longitudinal cognitive trajectories were assessed using CDR Sum of Boxes (CDR-SB) slopes, defined as the annual change in CDR-SB. Of the 700 participants recruited, 649 had CDR-SB recorded at baseline as well as at least one annual follow-up visit (mean ± SD follow-up duration, 50.5±15.9 months). These 649 participants were included in the calculation of CDR-SB slopes (no objective cognitive impairment, n = 132; cognitive impairment no dementia, n = 271; dementia, n = 246). To extract the CDR-SB slope for each participant, linear mixed-effects (LME) model modelling CDR-SB over time, with random intercepts and slopes, was used [21]. Next, incident cognitive decline was defined as an increase in CDR-GS of at least 0.5 at any annual follow-up visit compared to baseline [21].

### Neuroimaging biomarkers

Structural MRI was performed using a 3T Siemens Magnetom Trio Tim scanner (32-channel head coil). The protocol included 3D T1-weighted, T2-weighted, fluid-attenuated inversion recovery (FLAIR) and susceptibility-weighted imaging sequences. The following MRI biomarkers were assessed and included in the data analysis of the present study.

The MRI measures of neurodegeneration (“N”) include hippocampal volumes and medial temporal atrophy (MTA) scores. Brain volumes were measured from T1-weighted images using FreeSurfer v.6.0 (http://surfer.nmr.mgh.harvard.edu/). Briefly, the T1-weighted images were processed using standard and automated processing stream, including steps such as motion correction, intensity normalization, skull stripping, brain segmentation, tissue classification and volume calculation. Total hippocampal volume and intracranial volume (ICV) were extracted from the FreeSurfer output. Hippocampal volume was normalized to ICV and expressed as percentages (e.g. Hippocampal volume/ICV × 100%). MTA scores used in the sensitivity analyses were visually graded on coronal T1-weighted slices, according to the Scheltens scale.

The MRI measures of cerebral small vessel disease (“V”) include WMH volumes and Fazekas scores. WMHs were segmented using the Lesion Segmentation Toolbox (LST) v.3.0.0 with its lesion growth algorithm. WMH volume was then extracted for each patient, normalized to ICV, and expressed as percentages (e.g. WMH volume/ICV × 100%). Fazekas scores used in the sensitivity analyses were visually graded and defined as hyperintense on FLAIR sequences and hypointense on T1-weighted images, according to the modified Fazekas scale focusing on deep white matter.

### Plasma biomarkers

Non-fasting blood was collected into ethylenediaminetetraacetic acid (EDTA)– containing tubes and centrifuged at 2000g for 10 minutes at 4◦C. Plasma was extracted, mixed well, aliquoted and stored at -80◦C until use. Plasma samples were analyzed at Alamar Biosciences Inc. (Fremont, CA) using the NULISAseq™ CNS Disease Panel on the ARGO™ HT platform, with evaluators blinded to clinical information. Details have been published previously [5, 22]. Data were rescaled, added 1, and log2 transformed to obtain NULISA Protein Quantification (NPQ) units for downstream statistical analysis of BD-pTau and total-pTau species, including pTau217, pTau181, and pTau231, as well as GFAP and NfL. Selection of these biomarkers were based on established associations with amyloid pathology (“A”), inflammation (“I”), and neurodegeneration (“N”), respectively [5, 14, 23, 24]. Separately, a singleplex BD-pTau217 measurements used in the sensitivity analyses were derived using the NULISAqpcr BD-pTau217 assays (Alamar Biosciences), which provides absolute quantification of plasma BD-pTau217 concentration.

### Statistical Analyses

To determine the associations between MRI and plasma biomarkers with cognitive decline, participant-specific CDR-SB slopes derived from LME models were used as outcomes in linear regression models with individual MRI and plasma biomarkers as predictors (1 model per biomarker, adjusting for demographics, vascular risk factors, and baseline cognition). For comparison, we also fitted a basic model using only the covariates, without MRI and plasma biomarkers.

To identify the optimal biomarker combination for predicting CDR-SB slopes, we used the dredge function in the MuMIn package for R software to evaluate all combinations of MRI and plasma biomarkers while retaining the covariates in every model. The model with the lowest Akaike information criterion (AIC) was selected as the best-fitting model, representing the best tradeoff between fit and model complexity [25, 26]. A difference of 2 AICs is considered significant [25, 26]. Models within a difference of 2 AICs from the best-fitting model were scanned to identify the parsimonious model (e.g. model with the fewest predictors while maintaining similar performance)[25, 26]. We further assessed the performance of the best-fitting and parsimonious models in discriminating between individuals who experienced incident cognitive decline and those who remained cognitively stable at 2, 3, and 4 years. We used the area under the receiver-operating characteristic curve (AUC) and compared AUCs between these models and against the basic model using DeLong tests.

To assess the associations between the AVN groups and longitudinal cognitive performance, we used LME models with CDR-SB as outcome, including as predictors the AVN groups, time, an interaction between groups and time, demographics, vascular risk factors, and baseline cognition. The models contained random intercepts and random slopes, and time was modeled as a continuous variable. *Post-hoc* pairwise contrasts were conducted to compare the slopes for groups × time interaction. To enhance clinical interpretability, we converted the model-predicted annual CDR-SB change for each group into a model-implied years from 0.5 (within the normal range) to 6.5 (representing mid-range mild dementia) as (6.5 -0.5)/(ΔCDR-SB/year)[13].

To assess the associations between the AVN groups and risk of incident cognitive decline, we used Cox proportional hazards regression models, adjusting for demographics, vascular risk factors, and baseline cognition.

All analyses were performed using SPSS version 29 (IBM Corp., Armonk, NY) and R version 4.4.0 (R Project for Statistical Computing). Statistical significance was set at 2-tailed *P* < 0.05.

## Results

### Participant Characteristics

A total of 523 participants were included in the study (mean ± SD age, 72.4 ± 7.9 years; 291 females [55.6%]). At baseline, participants had no dementia or mild dementia, defined by a CDR-GS of 0 (n = 178 [34.0%]), 0.5 (n = 192 [36.7%]), or 1 (n = 153 [29.3%]). Measures of longitudinal cognition include the CDR-SB slope (change in CDR-SB per year, median [IQR], 0.24 [1.2]; mean ± SD, 0.67 ± 0.89; range, -0.38 to 4.36) and incident cognitive decline (increment in CDR-GS of ≥0.5 at annual follow-up compared with the baseline), over a mean ± SD follow-up time of 51.0 ± 15.8 months. Incident cognitive decline occurred in 226 participants (43.2%). Additional demographic, clinical, and biomarker information is shown in Table 1.

**Table 1.** Participant Demographic Characteristics, the Harmonisation cohort.

|  | **Harmonisation cohort (n =523)** |
|---|---|
| Age, mean (SD), y | 72.4 (7.9) |
| Female, No. (%) | 291 (55.6) |
| Education, mean (SD), y | 7.4 (5.1) |
| *APOE4* carrier, No. (%) | 154 (29.4) |
| Hypertension, No. (%) | 364 (69.6) |
| Hyperlipidemia, No. (%) | 382 (73.0) |
| Diabetes, No. (%) | 178 (34.0) |
| Cardiovascular disease, No. (%) | 61 (11.7) |
| Baseline MMSE, median (IQR) | 24 (7) |
| **Dementia severity** |  |
| CDR-GS = 0, No. (%) | 178 (34.0) |
| CDR-GS = 0.5, No. (%) | 192 (36.7) |
| CDR-GS = 1, No. (%) | 153 (29.3) |
| **Biomarkers** |  |
| Plasma BD-pTau217, mean (SD), NPQ<sup>a</sup> | 11.8 (0.7) |
| Plasma BD-pTau181, mean (SD), NPQ<sup>a</sup> | 13.9 (0.9) |
| Plasma BD-pTau231, mean (SD), NPQ<sup>a</sup> | 13.3 (0.3) |
| Plasma Total-pTau217, mean (SD), NPQ<sup>a</sup> | 11.4 (0.8) |
| Plasma Total-pTau181, mean (SD), NPQ<sup>a</sup> | 13.0 (0.8) |
| Plasma Total-pTau231, mean (SD), NPQ<sup>a</sup> | 13.3 (0.8) |
| Plasma NfL, mean (SD), NPQ<sup>a</sup> | 11.9 (0.9) |
| Plasma GFAP, mean (SD), NPQ<sup>a</sup> | 13.1 (0.8) |
| WMH volume, median (IQR), %<sup>b</sup> | 0.33 (0.7) |
| Hippocampal volume, mean (SD), %<sup>b</sup> | 0.48 (0.1) |
| **Longitudinal cognitive measures** |  |
| CDR-SB slope, median (IQR), change/y<sup>c</sup> | 0.24 (1.2) |
| CDR-SB slope, mean (SD), change/y<sup>c</sup> | 0.67 (0.9) |
| Incident cognitive decline, No. (%)<sup>d</sup> | 226 (43.2) |
| Follow-up time, months, mean (SD) | 51.0 (15.8) |

Abbreviations: *APOE4*, apolipoprotein E ε4 allele; BD, brain-derived; CDR-GS, clinical dementia rating global score; CDR-SB, clinical dementia rating sum of boxes; GFAP, glial fibrillary acidic protein; MMSE, Mini-Mental State Examination; NfL, neurofilament light; NPQ, NULISA protein quantification; WMH, white matter hyperintensity
<sup>a</sup> Blood samples used for plasma biomarker measurements were collected within 1.5 years of the baseline visit, with mean (SD) time from baseline visit = 2.7 (5) months
<sup>b</sup> WMH volume and hippocampal volume were normalized to intracranial volume as a percentage
<sup>c</sup> Participant-specific CDR-SB slopes were estimated from linear mixed-effects models with random intercepts and slopes
<sup>d</sup> Incident cognitive decline was defined as an increase in CDR-GS of ≥ 0.5 at annual follow-up compared with the baseline

### Combination of Plasma and MRI biomarkers for Predicting Cognitive Decline

In Table 2, adjusting for covariates, all plasma (BD-pTau217, BD-pTau181, BD-pTau231, total-pTau217, total-pTau181, total-pTau231, GFAP, and NfL) and MRI (WMH volume and hippocampal volume) biomarkers were significantly associated with CDR-SB slopes (all *P* < 0.001; adjusted R^2^ range, 0.381-0.477; AIC range, 1037-1125), yielding improved model fits as compared to the basic (covariates-only) model (adjusted R^2^, 0.367; AIC, 1136). Interestingly, all models with BD-pTau species showed better prediction performance than their respective total-pTau species (BD-pTau217 vs total-pTau217, adjusted R^2^ = 0.477 vs 0.454; ΔAIC = -22; BD-pTau181 vs total-pTau181, adjusted R^2^ = 0.458 vs 0.414; ΔAIC = -41; BD-pTau231 vs total-pTau231, adjusted R^2^ = 0.453 vs 0.431; ΔAIC = -21).

**Table 2.** Individual Biomarkers Associated With CDR-SB Slopes. ^a^

| **CDR-SB slopes<br>(n = 523)** | **β (95% CI)** | ***P* value** | **R<sup>2</sup>** | **Adjusted R<sup>2</sup>** | **AIC** |
|---|---|---|---|---|---|
| **Basic model (Covariates-only)** | NA | NA | 0.378 | 0.367 | 1135.756 |
| **Basic model + Plasma AD-related biomarker** |  |  |  |  |  |
| BD-pTau217 | 0.493 (0.400-0.586) | <.001 | 0.487 | 0.477 | 1036.861 |
| Total-pTau217 | 0.366 (0.287-0.444) | <.001 | 0.465 | 0.454 | 1058.864 |
| BD-pTau181 | 0.333 (0.263-0.403) | <.001 | 0.468 | 0.458 | 1055.287 |
| Total-pTau181 | 0.287 (0.200-0.374) | <.001 | 0.425 | 0.414 | 1096.154 |
| BD-pTau231 | 0.912 (0.715-1.110) | <.001 | 0.464 | 0.453 | 1059.930 |
| Total-pTau231 | 0.312 (0.233-0.392) | <.001 | 0.442 | 0.431 | 1080.500 |
| GFAP | 0.348 (0.254-0.441) | <.001 | 0.437 | 0.426 | 1085.535 |
| NfL | 0.184 (0.100-0.268) | <.001 | 0.399 | 0.388 | 1119.223 |
| **Basic model + MRI biomarker** |  |  |  |  |  |
| WMH volume | 0.176 (0.079-0.273) | <.001 | 0.393 | 0.381 | 1124.956 |
| Hippocampal volume | -2.508 (-3.462 to -1.555) | <.001 | 0.409 | 0.397 | 1111.170 |

Abbreviations: AD, Alzheimer's disease; AIC, Akaike information criterion; BD, brain-derived; CDR-SB, clinical dementia rating sum of boxes; GFAP, glial fibrillary acidic protein; MMSE, Mini-Mental State Examination; MRI, magnetic resonance imaging; NA, not applicable; NfL, neurofilament light; WMH, white matter hyperintensity.
<sup>a</sup> Participant-specific CDR-SB slopes, estimated from the linear mixed-effects model, were used as outcomes in separate linear regression models, with one model fitted for each biomarker. Each model was adjusted for age, sex, years of education, *APOE4* status, hypertension, hyperlipidemia, diabetes, and cardiovascular disease status, and baseline MMSE scores. For comparison, we also fitted a basic model using only the covariates, without biomarkers.

We next aimed to identify an optimal biomarker combination to predict CDR-SB slopes (Table 3). We evaluated all combinations of plasma and MRI biomarkers while retaining the covariates in every model. The best-fitting model for prediction of CDR-SB slopes (e.g. the model with the lowest AIC) included plasma BD-pTau217 (β ± SE = 0.391 ± 0.06; *P* < 0.001), plasma GFAP (β ± SE = 0.080 ± 0.06; *P* = 0.153), WMH volume (β ± SE = 0.139 ± 0.05; *P* = 0.002), and hippocampal volume (β ± SE = -1.30 ± 0.47; *P* = 0.006; AIC for the overall model = 1023; adjusted R^2^ = 0.493). The parsimonious model (e.g. model with the fewest predictors while maintaining a similar performance defined as being within two AIC points of the lowest AIC model) included plasma BD-pTau217 (β ± SE = 0.440 ± 0.05; *P* < 0.001), WMH volume (β ± SE = 0.142 ± 0.05; *P* = 0.002), and hippocampal volume (β ± SE = -1.35 ± 0.47; *P* = 0.004; AIC for the overall model = 1023; adjusted R^2^ = 0.492). We also tested the three-way interaction amongst the three biomarkers as well as each corresponding two-way interaction, none of which was significant (all *P* > 0.10).

**Table 3.** Model Selection Process and Performance of Each Model for Identifying Incident Cognitive Decline.

| Linear regression model selection process<sup>a</sup><br>(Outcome: CDR-SB slopes)<br>(n = 523) |  |  | Performance of each model for identifying incident cognitive decline<sup>b</sup><br>(Outcome: Increment in CDR-GS of ≥ 0.5 at follow-up) |  |  |
|---|---|---|---|---|---|
| Model | Variables | AIC | AUC<br>(Within 2 years)<br>Stable: 354 (72%)<br>Decline: 135 (28%) | AUC<br>(Within 3 years)<br>Stable: 295 (63%)<br>Decline: 174 (37%) | AUC<br>(Within 4 years)<br>Stable: 247 (55%)<br>Decline: 203 (45%) |
| **Best-fitting model** | Basic model + plasma BD-pTau217 + plasma GFAP + WMH volume + Hippocampal volume | 1023.222 | 0.768 (0.720-0.816)<sup>c</sup> | 0.805 (0.762-0.847)<sup>d</sup> | 0.825 (0.785, 0.864)<sup>d</sup> |
| **Parsimonious model** | Basic model + plasma BD-pTau217 + WMH volume + Hippocampal volume | 1023.326 | 0.768 (0.719-0.816)<sup>c</sup> | 0.805 (0.762-0.847)<sup>d</sup> | 0.823 (0.784, 0.863)<sup>d</sup> |
| **Basic model (Covariates-only)** | Age, sex, years of education, *APOE4* status, hypertension, hyperlipidemia, diabetes, and cardiovascular disease status, and baseline MMSE scores | 1135.756 | 0.740 (0.690-0.789) | 0.763 (0.718-0.809) | 0.779 (0.736, 0.823) |

Abbreviations: AIC, Akaike information criterion; AUC, area under the ROC curve; BD, brain-derived; CDR-GS, clinical dementia rating global score; CDR-SB, clinical dementia rating sum of boxes; GFAP, glial fibrillary acidic protein; MMSE, Mini-Mental State Examination; ROC, receiver operating characteristic; WMH, white matter hyperintensity.
<sup>a</sup> The linear regression model selection process. Best-fitting model shows the data-driven model selection with the lowest AIC. The parsimonious model shows the model that had a similar model performance (ΔAIC < 2) with as few predictors as possible.
<sup>b</sup> ROC curve analyses of the different models for discriminating individuals with incident cognitive decline versus those who remained cognitively stable, within 2, 3, or 4 years. Incident cognitive decline was defined as an increase in CDR-GS of ≥0.5 at annual follow-up compared with the baseline. Within each time period, comparisons of AUCs were performed using DeLong test.
<sup>c</sup> *P*□≤□0.02 compared with the basic model
<sup>d</sup> *P*□≤□0.001 compared with the basic model

We further assessed the performance of the best-fitting and parsimonious models in discriminating between individuals who experienced incident cognitive decline and those who remained cognitively stable at 2, 3, and 4 years (Table 3). Both models demonstrated similar accuracy (best-fitting model vs parsimonious model; AUC [95% CI], 2 years: 0.768 [0.720-0.816] vs 0.768 [0.719-0.816]; 3 years: 0.805 [0.762-0.847] vs 0.805 [0.762-0.847]; 4 years: 0.825 [0.785-0.864] vs 0.823 [0.784-0.863]). Both models significantly outperformed the basic model (AUC [95% CI], 2 years: 0.740 [0.690-0.789]; both *P* ≤ 0.02; 3 years: 0.763 [0.718-0.809]; both *P* ≤ 0.001; 4 years: 0.779 [0.736-0.823]; both *P* ≤ 0.001).

### Association between Increasing Number of Positive AVN Biomarkers and Cognitive Decline

Given the similar performance of the best-fitting and parsimonious models in predicting cognitive decline, subsequent analyses focused on the biomarkers selected in the parsimonious model, including plasma BD-pTau217 (associated with amyloid pathology; “A”), WMH volume (associated with cerebral small vessel disease; “V”), and hippocampal volume (associated with neurodegeneration; “N”).

To assess the association between increasing number of positive AVN biomarkers and cognitive decline, each biomarker was dichotomized to derive A, V, and N status (see Supplemental Table 1 for positivity thresholds). As shown in Supplemental Table 2, participants who were negative for all three biomarkers (A-V-N-; n = 128) were classified as “all negative”. Participants who were positive for one biomarker (A+V-N-, A-V+N-, or A-V-N+; n = 154) were collectively classified as “1-positive”. Participants who were positive for two biomarkers (A+V+N-, A+V-N+, or A-V+N+; n = 158) were collectively classified as “2-positive”. Participants who were positive for all three biomarkers (A+V+N+; n = 83) were classified as “3-positive”.

Annual CDR-SB progression increased stepwise with the number of positive biomarkers (Figure 2). Compared with the All negative group (β [95% CI], 0.016 [-0.135 to 0.168]), participants in the 1-positive (β [95% CI], 0.409 [0.268-0.549]), 2-positive (β [95% CI], 1.16 [1.02-1.30]) and 3-positive groups (β [95% CI], 1.49 [1.29-1.69]) showed faster cognitive decline (all *P* < 0.001). Moreover, the 2-positive and 3-positive groups progressed faster than the 1-positive group (both *P* < 0.001), and the 3-positive group declined faster than 2-positive group (*P* = 0.009). Based on the model-predicted annual CDR-SB changes, participants in the 1-positive group were model-implied to take approximately 15 years to progress from a CDR-SB of 0.5 to 6.5. This window shortened with more positive biomarkers: approximately 5 years for the 2-positive group, and 4 years for the 3-positive group. These estimates are translations of predicted slopes, not time-to-event estimates. Overall, these findings illustrate that a greater number of positive pathological biomarkers is associated with accelerated clinical progression.

Similar results were observed for incident cognitive decline (Table 4). The incidence of cognitive decline was 14% (18/128), 36% (55/154), 61% (96/158), and 69% (57/83) in the all negative, 1-positive, 2-positive, and 3-positive groups, respectively. Cox regression showed that the 1-positive, 2-positive, and 3-positive groups each had a higher risk of incident cognitive decline than the all negative group (all *P* < 0.001). Risk increased with the number of positive biomarkers and was highest in the 3-positive group (hazard ratio [95% CI], 1-positive: 2.93 [1.69-5.07]; 2-positive: 6.20 [3.54-10.8]; 3-positive: 7.15 [3.91-13.1]).

**Table 4.**
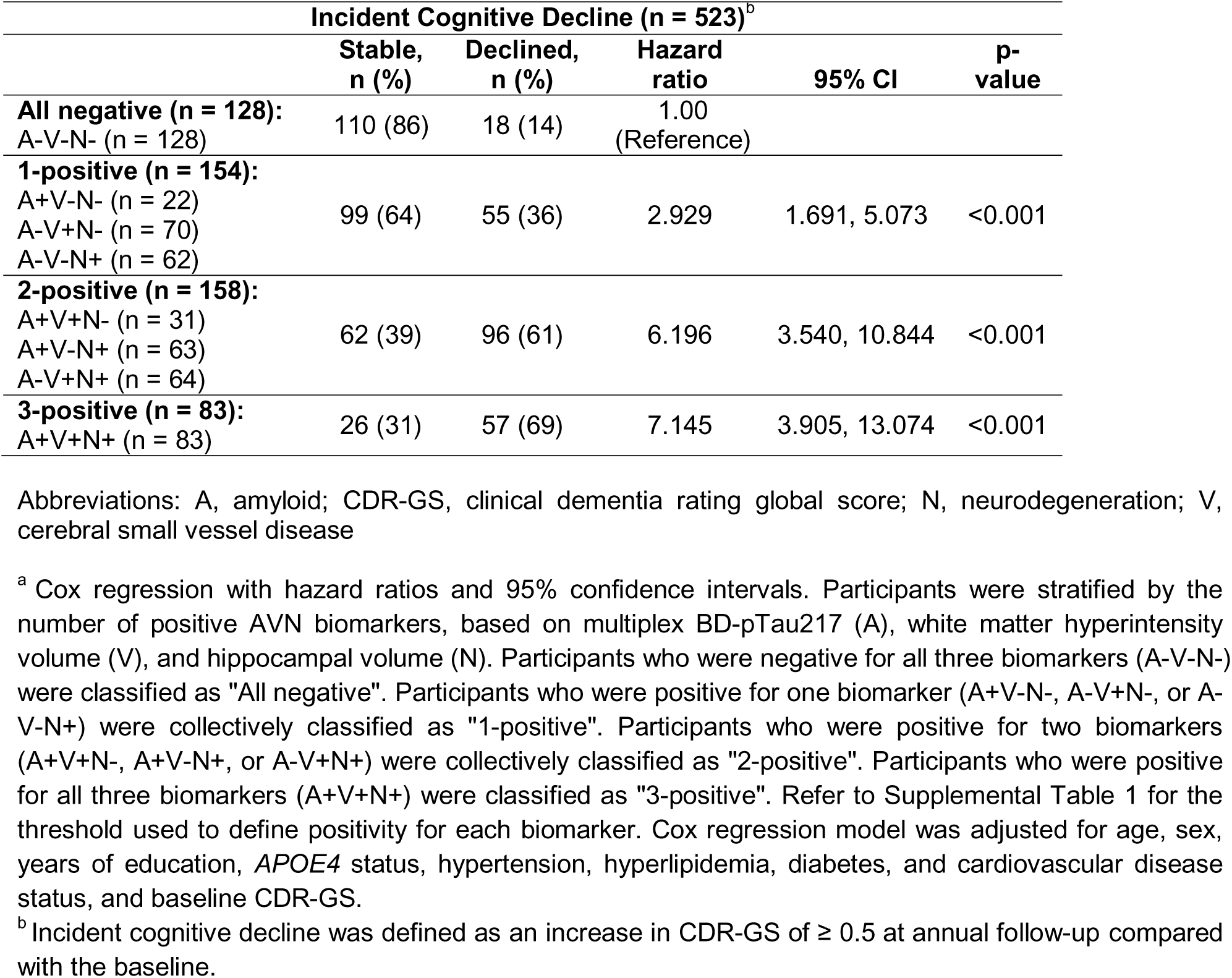
Cox Regression Analyses of Risk for Incident Cognitive Decline in Participants Stratified by Number of Positive AVN Biomarkers^a^.

| Incident Cognitive Decline (n = 523)<sup>b</sup> |  |  |  |  |  |
|---|---|---|---|---|---|
|  | **Stable, n (%)** | **Declined, n (%)** | **Hazard ratio** | **95% CI** | **p-value** |
| **All negative (n = 128):**<br>A-V-N- (n = 128) | 110 (86) | 18 (14) | 1.00 (Reference) |  |  |
| **1-positive (n = 154):**<br>A+V-N- (n = 22)<br>A-V+N- (n = 70)<br>A-V-N+ (n = 62) | 99 (64) | 55 (36) | 2.929 | 1.691, 5.073 | <0.001 |
| **2-positive (n = 158):**<br>A+V+N- (n = 31)<br>A+V-N+ (n = 63)<br>A-V+N+ (n = 64) | 62 (39) | 96 (61) | 6.196 | 3.540, 10.844 | <0.001 |
| **3-positive (n = 83):**<br>A+V+N+ (n = 83) | 26 (31) | 57 (69) | 7.145 | 3.905, 13.074 | <0.001 |

Abbreviations: A, amyloid; CDR-GS, clinical dementia rating global score; N, neurodegeneration; V, cerebral small vessel disease
<sup>a</sup> Cox regression with hazard ratios and 95% confidence intervals. Participants were stratified by the number of positive AVN biomarkers, based on multiplex BD-pTau217 (A), white matter hyperintensity volume (V), and hippocampal volume (N). Participants who were negative for all three biomarkers (A-V-N-) were classified as "All negative". Participants who were positive for one biomarker (A+V-N-, A-V+N-, or A-V-N+) were collectively classified as "1-positive". Participants who were positive for two biomarkers (A+V+N-, A+V-N+, or A-V+N+) were collectively classified as "2-positive". Participants who were positive for all three biomarkers (A+V+N+) were classified as "3-positive". Refer to Supplemental Table 1 for the threshold used to define positivity for each biomarker. Cox regression model was adjusted for age, sex, years of education, *APOE4* status, hypertension, hyperlipidemia, diabetes, and cardiovascular disease status, and baseline CDR-GS.
<sup>b</sup> Incident cognitive decline was defined as an increase in CDR-GS of ≥ 0.5 at annual follow-up compared with the baseline.

### Sensitivity Analyses

Two sets of sensitivity analyses were performed. First, to improve feasibility for clinical implementation, we repeated the AVN analyses using plasma BD-pTau217 measured with a singleplex assay that provided absolute quantification of this biomarker (“A”), visual ratings of Fazekas scores (“V”) and medial temporal atrophy scores (“N”; see Supplemental Tables 1 and 2), with similar results observed (Supplemental Figure 2 and Supplemental Table 3). Compared with the All negative group (β [95% CI], 0.096 [-0.063 to 0.256]), participants in the 1-positive (β [95% CI], 0.496 [0.350-0.641]), 2-positive (β [95% CI], 1.03 [0.881-1.18]) and 3-positive groups (β [95% CI], 1.53 [1.31-1.74]) showed faster cognitive decline (all *P* < 0.001). The 2-positive and 3-positive groups progressed faster than the 1-positive group (both *P* < 0.001), and the 3-positive group declined faster than 2-positive group (*P* < 0.001). Based on the model-predicted annual CDR-SB changes, participants in the 1-positive group were model-implied to take approximately 12 years to progress from a CDR-SB of 0.5 to 6.5. This window shortened with more positive biomarkers: approximately 6 years for the 2-positive group, and 4 years for the 3-positive group.

The incidence of cognitive decline was 18% (22/126), 39% (62/158), 57% (89/155), and 64% (51/80) in the all negative, 1-positive, 2-positive, and 3-positive groups, respectively. Risk increased with the number of positive biomarkers (hazard ratio [95% CI], 1-positive: 2.48 [1.50-4.10]; 2-positive: 3.88 [2.32-6.49]; 3-positive: 4.74 [2.66-8.47]; all *P* < 0.001).

Second, we assessed the longitudinal CDR-SB trajectories across the eight AVN combinations (A-V-N-, A+V-N-, A-V+N-, A-V-N+, A+V+N-, A+V-N+, A-V+N+, A+V+N+). Using the multiplex plasma BD-pTau217 (“A”), WMH volume (“V”), and hippocampal volume (“N”), we observed a generally increasing rate of annual CDR-progression with greater biomarker positivity. The estimated β values, from lowest to highest: A-V-N-(all negative), 0.017; A-V+N-(1-positive), 0.382; A-V-N+ (1-positive), 0.410; A+V-N-(1-positive), 0.477; A+V+N-(2-positive), 0.857; A-V+N+ (2-positive), 0.914; A+V+N+ (3-positive), 1.49; A+V-N+ (2-positive), 1.54 (Supplemental Figure 3A). Cox regression analyses yielded similar observations (Supplemental Table 4A). Briefly, the hazard ratios, from lowest to highest: A-V-N-(all negative; reference group), 1.00; A+V-N-(1-positive), 2.36; A-V-N+ (1-positive), 2.52; A-V+N-(1-positive), 3.30; A-V+N+ (2-positive), 4.11; A+V+N-(2-positive), 5.37; A+V+N+ (3-positive), 7.12; A+V-N+ (2-positive), 9.14. Results based on singleplex plasma BD-pTau217 (“A”), Fazekas scores (“V”), and medial temporal atrophy scores (“N”) are shown in Supplemental Figure 3B and Supplemental Table 4B.

## Discussion

In this large Southeast Asian cohort that is characterized by a high prevalence of CeVD, we identified an optimal multimodal biomarker combination for predicting cognitive decline and clinical progression: plasma BD-pTau217, WMH volume, and hippocampal volume, reflecting amyloid pathology (“A”), CeVD (“V”), and neurodegeneration (“N”), respectively. All three biomarkers were significantly and independently associated with faster cognitive decline, as measured by CDR-SB slopes. Together, they achieved high performance in discriminating individuals who experienced clinical progression, defined as an increase in CDR-GS of at least 0.5 at follow-up, from those who remained cognitively stable. To further examine the association between positivity for multiple pathological biomarkers and cognitive decline, we stratified participants by the number of positive biomarkers. We observed a stepwise increase in the rate of cognitive decline as the number of positive biomarkers increased. Similarly, the risk of clinical progression increased with the number of positive biomarkers. Remarkably, among participants who were positive for all three biomarkers (A+V+N+), approximately 70% experienced progression during the maximum follow-up period of five years. The hazard of progression was approximately seven times higher in this group than in the reference group (A−V−N−; HR = 7.15). Overall, these results highlight the utility of a multimodal biomarker profile that captures the key neuropathological features and biological processes driving cognitive decline to improve prognostication in a given cohort. They support similar assessments in other cohorts, which may be characterized by different neuropathological burden and biologically important processes.

Consistent with previous studies, each plasma and MRI biomarker was individually associated with faster cognitive decline and increased risk of clinical progression[2, 5, 27–31], supporting a potential link between these pathologies and biological processes and cognitive deterioration. When all biomarkers were considered in the model selection process, only plasma BD-pTau217, WMH volume, and hippocampal volume were selected. This combination reflects the predominant pathological burden in this cohort, namely AD pathology and CeVD, as well as nonspecific neurodegeneration beyond that attributable to AD and CeVD. The utility of this multimodal prognostic model is further supported by our observation that groups positive for at least two AVN biomarkers showed faster cognitive decline and a higher risk of clinical progression than those with one or no positive biomarkers. A few findings warrant further discussion. First, all BD-pTau species outperformed their respective total pTau species in predicting cognitive decline. This could be related to our previous observation that BD-pTau species showed greater accuracy for identifying amyloid burden than total-pTau species[5, 6], which may translate to better prediction of AD-related cognitive decline. Second, although both plasma NfL and hippocampal volume were considered biomarkers of neurodegeneration, hippocampal volume was selected for the model. One possible explanation is that elevated plasma NfL reflects neuroaxonal injury across non-specific neurodegenerative diseases and brain regions[32, 33], including those less directly implicated in cognitive performance, whereas reduced hippocampal volume reflects neurodegeneration in a brain region that plays a major role in memory[34]. Third, the exclusion of plasma GFAP from the parsimonious model does not indicate that inflammation is irrelevant to cognitive deterioration in our cohort. Although elevated GFAP is a strong indication of reactive astrocyte remodeling, it is not an absolute or sole marker of reactivity[35]. Future work should consider other markers of reactive astrocytes, such as YKL40 and S100B[35], as well as markers of microglial activation, inflammatory cytokines and chemokines[36]. Finally, our results also have implications for current disease-targeting treatments (DTTs) directed at a single pathology, such as amyloid. Because multiple pathologies may contribute to accelerated cognitive decline, coexisting pathologies could partly explain the variable or reduced overall efficacy observed with such treatments. For example, even if amyloid is completely removed from the brain, substantial cognitive decline may still occur in the presence of coexisting pathologies.

There are limitations to this study: first, other non-AD pathologies, such as α-synucleinopathies and limbic-predominant age-related TDP-43 encephalopathy (LATE), were not assessed in this study due to the lack of biomarkers. Such pathologies could partly explain the findings in A−V−N+ participants, who showed evidence of significant neurodegeneration despite low amyloid and WMH burden. Second, the presence of both amyloid and tau pathology has previously been associated with faster cognitive decline[37]. Adding a tau biomarker may therefore further improve the current prognostic model. It could also help determine whether significant neurodegeneration in A+V−N+ individuals occur alongside established tau pathology, consistent with a later biological stage of AD (e.g., A+T+), or in the absence of substantial tau pathology (e.g., A+T-), potentially suggesting the contribution of coexisting non-AD pathologies. Future studies could include emerging plasma biomarkers of aggregated and established tau burden, such as eMTBR-tau[38]. Third, our prognostic model was derived from a single cohort. In addition to validating the current AVN model in diverse cohorts, future studies should conduct similar analyses in cohorts with different neuropathological burden. These analyses could help develop prognostic models optimized for cohorts that closely resemble their intended-use populations. Finally, given previous observations that some individuals remain cognitively stable despite a high neuropathological burden, future studies should examine resilience factors and biomarkers alongside neuropathological biomarkers associated with cognitive decline [21, 39–41]. These resilience factors and biomarkers may help explain the heterogeneity in cognitive trajectories and further enhance prognostication.

In conclusion, the presence of multiple neuropathophysiological processes is associated with accelerated cognitive decline. A multimodal model incorporating biomarkers of amyloid pathology, CeVD, and neurodegeneration was the optimal combination for predicting cognitive decline in this cohort with a high CeVD burden (Figure 1), although it may be refined by incorporating the additional biomarkers discussed earlier. Future studies should aim to derive and optimize multimodal prognostic models in cohorts that resemble their intended-use populations.

**Figure 1.**
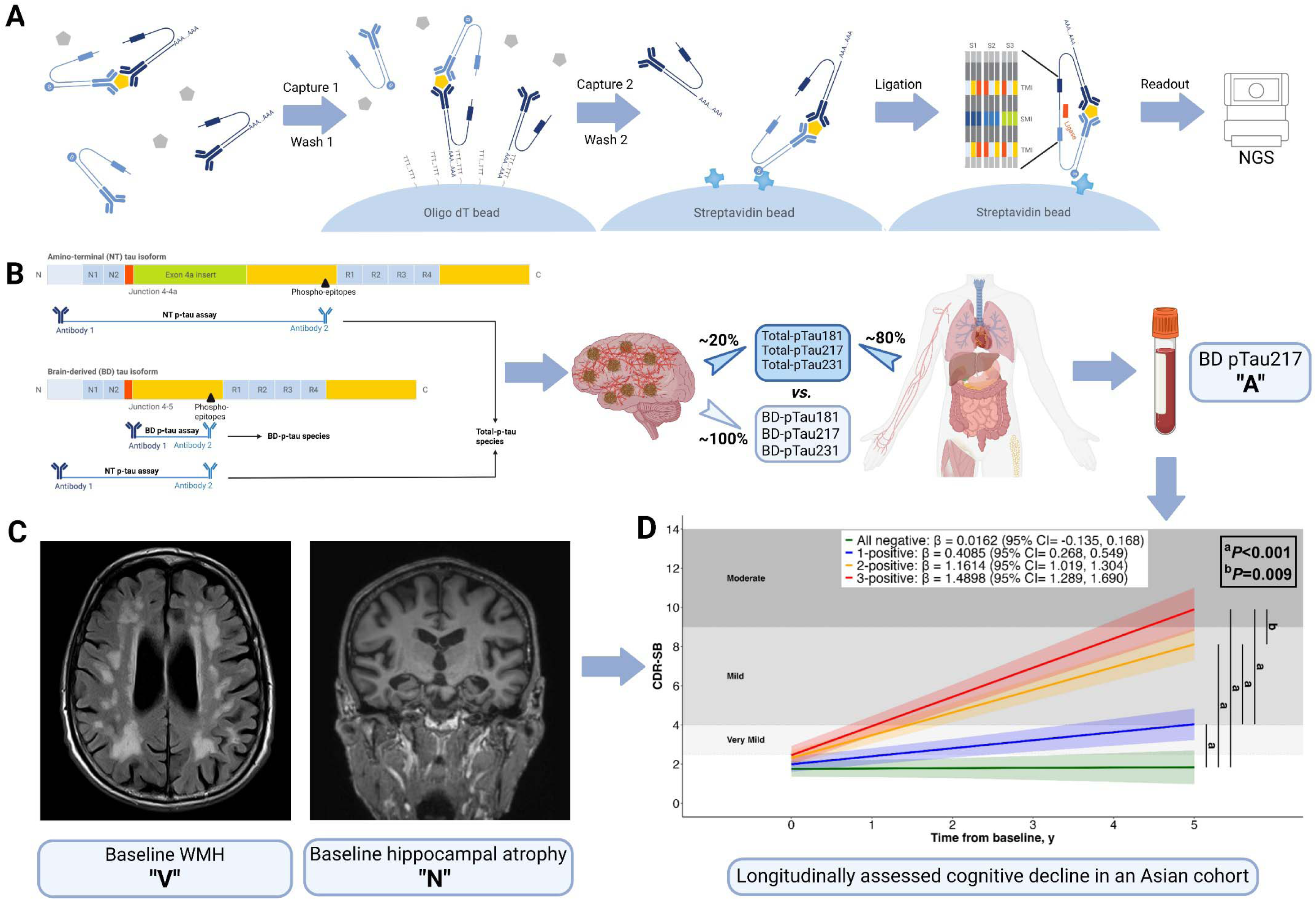
Graphical overview of study workflow and major findings. (A) Principle of NULISAseq, consisting of DNA-barcode-containing detection-capture antibody pairs, one conjugated with poly-adenine nucleotides and the other with biotin. Each pair of antibodies recognize different epitopes on the same target molecule, forming a tripartite immune complex, which are then sequentially captured and washed on oligo-dT beads (“Capture 1 / Wash 1”), and subsequently on streptavidin beads (“Capture 2 / Wash 2”), with the proximal ends of the DNA strands of the retained, complexed antibodies ligated to generate a DNA reporter molecule containing both target-specific and sample-specific barcodes. DNA reporter molecules were pooled and amplified by polymerase chain reaction (PCR), purified and sequenced on NGS, with sequencing data processed by NULISAseq algorithm for the quantification of sample-(SMI) and target-specific (TMI) barcodes (molecular identifiers) to derive NULISA Protein Quantification (NPQ) units for downstream statistical analysis. (B) Specificity for BD (LMW) Tau isoforms was achieved with antibodies which recognize an epitope at the junction between MAPT exons 4 and 5, while antibodies which recognize the common amino terminus (NT) region will detect both LMW and HMW isoforms. These antibodies are paired with antibodies specific for phospho-epitopes at threonine-181, 217 or 231, thus enabling the measurements of BD- and total-p-Tau species, with postulated proportions from brain and peripheral organ sources listed. Of the 6 pTau species investigated, BD-pTau217 performed best as the marker for “A”. (C) Neuroimaging biomarkers for “V” and “N” based on baseline WMH and baseline hippocampal atrophy. (D) The plasma and neuroimaging panel for AVN incrementally contribute to longitudinal cognitive impairment. Abbreviations: BD, Brain-derived; CDR-SB, Clinical Dementia Rating Sum of Boxes; NGS, Next-Generation sequencer; NT, Amino terminal; NULISAseq, NUcleic acid Linked Immuno-Sandwich Assay multiplexing platform; SMI, Sample-specific molecular identifier; TMI, Target-specific molecular identifier; WMH, white matter hyperintensities.

**Figure 2.**
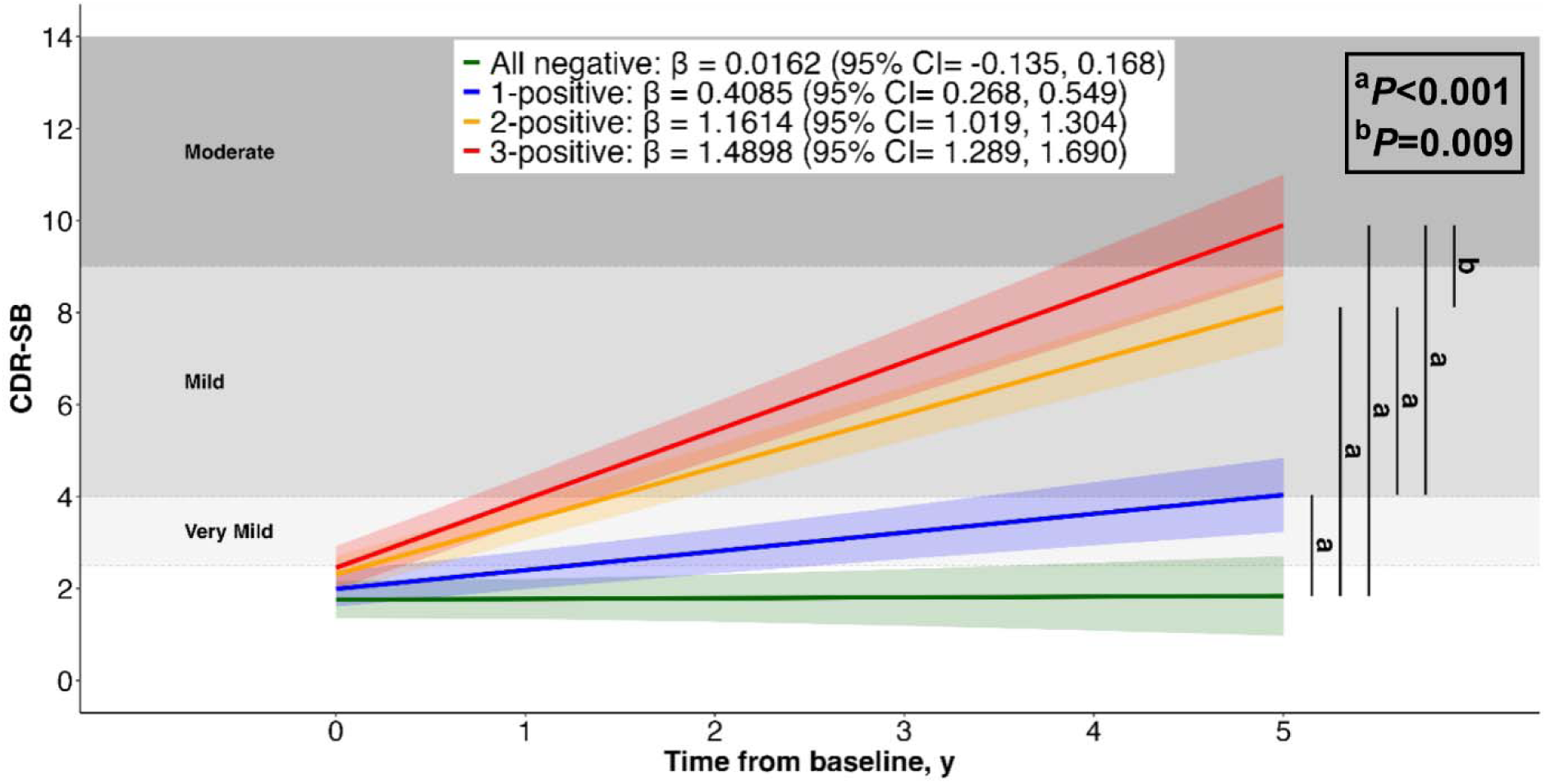
Longitudinal Trajectories of Clinical Dementia Rating Sum of Boxes According to Number of Positive AVN Biomarkers. Figure 1. Trajectory plots indicate the mean longitudinal trajectories (solid line) of clinical dementia rating sum of boxes (CDR-SB), and associated 95% confidence intervals (shading), estimated with linear mixed-effects model. Trajectories are stratified based on the number of positive AVN biomarkers (All negative [green], 1-positive [blue], 2-positive [orange], 3-positive [red]), modelled with an interaction between group and time. Participants were stratified based on multiplex BD-pTau217 (A), white matter hyperintensity volume (V), and hippocampal volume (N). Participants who were negative for all three biomarkers (A-V-N-) were classified as “All negative”. Participants who were positive for one biomarker (A+V-N-, A-V+N-, or A-V-N+) were collectively classified as “1-positive”. Participants who were positive for two biomarkers (A+V+N-, A+V-N+, or A-V+N+) were collectively classified as “2-positive”. Participants who were positive for all three biomarkers (A+V+N+) were classified as “3-positive”. Refer to Supplemental Table 1 for the threshold used to define positivity for each biomarker. Model included random slopes and intercepts and was adjusted for age, sex, years of education, *APOE4* status, hypertension, hyperlipidemia, diabetes, and cardiovascular disease status, and baseline clinical dementia rating global score (CDR-GS). Shaded background regions indicate dementia severity categories based on CDR-SB thresholds[42]: 2.5–4.0 (very mild), 4.5–9.0 (mild), 9.5–15.5 (moderate), and 16.0–18.0 (severe). *P* values represent *post-hoc* pairwise comparing the slopes between groups.

## Acknowledgments

We would like to thank the study patients and their caregivers for their support. The Graphical Abstract was created in BioRender. This study was supported by the Technology Access Program, Alamar Biosciences, Inc. which played no role in the writing of this manuscript or the decision to publish.

## CRediT authorship contribution statement

**Joyce Chong:** Conceptualization, Methodology, Formal analysis, Validation, Data curation, Writing – Original Draft. **Saima Hilal:** Investigation, Resources, Writing – Review & Editing. **Thomas Karikari:** Resources, Writing – Review & Editing. **Narayanaswamy Venketasubramanian:** Investigation, Resources, Writing – Review & Editing. **Michael Schöll:** Resources, Writing – Review & Editing. **Henrik Zetterberg:** Resources, Writing – Review & Editing. **Sterling Johnson:** Resources, Writing – Review & Editing. **Christopher Chen:** Investigation, Resources, Methodology, Supervision, Funding acquisition, Writing – Review & Editing. **Mitchell Lai:** Conceptualization, Validation, Data curation, Supervision, Funding acquisition, Writing – Original Draft.

## Data availability

Data is available from the corresponding authors upon reasonable request.

## Declarations

### Ethics approval and consent to participate

Study approval was obtained from Singapore National Healthcare Group Domain-Specific Review Board (2018/00996, 2015/00406, and 2015/00441). Written informed consent was obtained from study participants or their next-of-kin.

### Supplementary information

• Supplemental Figure 1. Flow Chart of Study Participants n = 523

• Supplemental Figure 2. Longitudinal Trajectories of Clinical Dementia Rating Sum of Boxes According to Number of Positive AVN Biomarkers (Based on Singleplex BD-pTau217, Fazekas score, and Medial Temporal Atrophy Score)

• Supplemental Figure 3. Longitudinal Trajectories of Clinical Dementia Rating Sum of Boxes According to AVN Biomarkers

• Supplemental Table 1. Dichotomization of Plasma BD-pTau217, White Matter Hyperintensity and Hippocampal volumes

• Supplemental Table 2. Stratification of Participants into AVN Groupings

• Supplemental Table 3. Cox Regression Analyses of Risk for Incident Cognitive Decline in Participants Stratified by Number of Positive AVN Biomarkers (Based on Singleplex BD-pTau217, Fazekas score, and Medial Temporal Atrophy Score)

• Supplemental Table 4. Cox Regression Analyses of Risk for Incident Cognitive Decline in Participants Stratified by AVN Biomarkers

## Abbreviations

“A”: Amyloid
AD: Alzheimer’s disease
AIC: Akaike information criterion
*APOE4*: Apolipoprotein E ε4 allele
AUC: Area under the receiver-operating characteristic curve
AVN: Amyloid, cerebral small vessel disease (vascular), neurodegeneration
BD: Brain-derived
CDR: Clinical Dementia Rating
CDR-GS: Clinical Dementia Rating Global Score
CDR-SB: Clinical Dementia Rating Sum of Boxes
CeVD: Cerebrovascular disease
CI: Confidence interval
EDTA: Ethylenediaminetetraacetic acid
FLAIR: Fluid-attenuated inversion recovery
GFAP: Glial fibrillary acidic protein
HR: Hazard ratio
“I”: Inflammation
ICV: Intracranial volume
IQR: Interquartile range
LATE: Limbic-predominant age-related TDP-43 encephalopathy
LME: Linear mixed-effects
LST: Lesion Segmentation Toolbox
MMSE: Mini-Mental State Examination
MRI: Magnetic resonance imaging
MTA: Medial temporal atrophy
“N”: Neurodegeneration
NfL: Neurofilament light chain
NPQ: NULISA Protein Quantification
PET: Positron emission tomography
pTau: Phosphorylated tau
ROC: Receiver operating characteristic
“S”: α-synuclein
SD: Standard deviation
SE: Standard error
“V”: Cerebral small vessel disease (vascular)
WMH: White matter hyperintensity

## References

1. Therriault, J., et al., Blood phosphorylated tau for the diagnosis of Alzheimer’s disease: a systematic review and meta-analysis. Lancet Neurol, 2025. 24(9): p. 740–752.

2. Chong, J.R., et al., Clinical utility of plasma p-tau217 in identifying abnormal brain amyloid burden in an Asian cohort with high prevalence of concomitant cerebrovascular disease. Alzheimers Dement, 2025. 21(2): p. e14502.

3. Buckley, R.F., et al., Prognostic Value of Blood-Based P-Tau217 Levels for Progression to Cognitive Impairment. Jama, 2026. 336(11): p. 950–959.

4. Lehmann, S., et al., *Clinical value of plasma ALZpath pTau217 immunoassay for assessing mild cognitive impairment.* Journal of Neurology, Neurosurgery & Psychiatry, 2024. 95(11): p. 1046.

5. Chong, J.R., et al., Plasma brain-derived p-Tau217 outperforms other p-Tau species in detecting abnormal brain amyloid in an Asian cohort of older people with cerebrovascular disease burden. The Journal of Prevention of Alzheimer’s Disease, 2026. 13(7): p. 100615.

6. Jiang, Y., et al., Head-to-head comparison of brain-derived pTau217 and total pTau217 for brain amyloid and tau pathology classification. Proc Natl Acad Sci U S A, 2026. 123(10): p. e2536792123.

7. Robinson, J.L., et al., Neurodegenerative disease concomitant proteinopathies are prevalent, age-related and APOE4-associated. Brain, 2018. 141(7): p. 2181–2193.

8. McAleese, K.E., et al., Concomitant neurodegenerative pathologies contribute to the transition from mild cognitive impairment to dementia. Alzheimers Dement, 2021. 17(7): p. 1121–1133.

9. Nichols, E., et al., The prevalence, correlation, and co-occurrence of neuropathology in old age: harmonisation of 12 measures across six community-based autopsy studies of dementia. Lancet Healthy Longev, 2023. 4(3): p. e115–e125.

10. Lam, B.Y.K., et al., High burden of cerebral white matter lesion in 9 Asian cities. Sci Rep, 2021. 11(1): p. 11587.

11. Lam, B.Y.K., et al., Global burden of cerebral small vessel disease determined from large MRI studies: A systematic review and meta-analysis. Int J Stroke, 2026. 21(9): p. 1284–1298.

12. Palmqvist, S., et al., Cognitive effects of Lewy body pathology in clinically unimpaired individuals. Nature Medicine, 2023. 29(8): p. 1971–1978.

13. Du, L., et al., Tau mediates the impact of amyloid and vascular disease burden on the trajectory of clinical symptoms. Alzheimers Dement, 2025. 21(10): p. e70831.

14. Jack, C.R., Jr., et al., Revised criteria for diagnosis and staging of Alzheimer’s disease: Alzheimer’s Association Workgroup. Alzheimers Dement, 2024. 20(8): p. 5143–5169.

15. Collij, L.E., et al., Lewy body pathology exacerbates brain hypometabolism and cognitive decline in Alzheimer’s disease. Nat Commun, 2024. 15(1): p. 8061.

16. Yang, J., et al., Interactive Effects of Plasma Alzheimer Disease Biomarkers and White Matter Hyperintensities on Cognitive Decline and Clinical Progression in Nondemented Adults. J Am Heart Assoc, 2026. 15(16): p. e050676.

17. Wu, C.Y., et al., Combined use of plasma p-tau217, NfL, and GFAP predicts domain-specific cognitive decline in cognitively unimpaired and MCI individuals. Alzheimers Dement, 2025. 21(12): p. e70934.

18. Bachmann, D., et al., Independent and interactive contributions of white matter hyperintensities and Alzheimer’s disease imaging and plasma biomarkers to cognitive decline in older adults without dementia. Alzheimer’s Research & Therapy, 2026. 18(1): p. 106.

19. Bellaver, B., et al., Astrocyte reactivity identifies p-tau217-positive individuals at elevated risk of cognitive decline across the Alzheimer’s disease continuum. Molecular Neurodegeneration, 2026.

20. Lim, M.J., et al., HARMONISATION – A multimodal prospective study of vascular cognitive impairment in multi-ethnic Asians: Cohort profile, progress, current contributions, and future impact. Journal of Alzheimer’s Disease. 0(0): p. 13872877251389006.

21. Chong, J.R., et al., Metabolism of the antioxidant micronutrient ergothioneine as a plasma biomarker of cognitive resilience in older people with Alzheimer’s disease amyloid pathology. Free Radic Biol Med, 2026. 254: p. 370–377.

22. Feng, W., et al., NULISA: a proteomic liquid biopsy platform with attomolar sensitivity and high multiplexing. Nature Communications, 2023. 14(1): p. 7238.

23. Chong, J.R., et al., Association of plasma GFAP with elevated brain amyloid is dependent on severity of white matter lesions in an Asian cognitively impaired cohort. Alzheimers Dement (Amst), 2024. 16(2): p. e12576.

24. Chong, J.R., et al., Brain atrophy and white matter hyperintensities are independently associated with plasma neurofilament light chain in an Asian cohort of cognitively impaired patients with concomitant cerebral small vessel disease. Alzheimers Dement (Amst), 2023. 15(1): p. e12396.

25. Mattsson-Carlgren, N., et al., Prediction of Longitudinal Cognitive Decline in Preclinical Alzheimer Disease Using Plasma Biomarkers. JAMA Neurol, 2023. 80(4): p. 360–369.

26. Palmqvist, S., et al., Prediction of future Alzheimer’s disease dementia using plasma phospho-tau combined with other accessible measures. Nat Med, 2021. 27(6): p. 1034– 1042.

27. Mielke, M.M., et al., Plasma and CSF neurofilament light: Relation to longitudinal neuroimaging and cognitive measures. Neurology, 2019. 93(3): p. e252–e260.

28. Ghanbarian, E., et al., Prognostic value of plasma glial fibrillary acidic protein in cognitively unimpaired older adults: Results from the A4 study. Alzheimers Dement, 2025. 21(11): p. e70948.

29. Cetindag, A.C., et al., Plasma GFAP outperforms CSF GFAP in detecting amyloid pathology and is associated with increased risk of clinical progression in early Alzheimer’s disease. J Prev Alzheimers Dis, 2026. 13(5): p. 100544.

30. Mattsson, N., et al., Association of Plasma Neurofilament Light With Neurodegeneration in Patients With Alzheimer Disease. JAMA Neurol, 2017. 74(5): p. 557–566.

31. Ding, D., et al., White Matter Hyperintensity Predicts the Risk of Incident Cognitive Decline in Community Dwelling Elderly. J Alzheimers Dis, 2018. 61(4): p. 1333–1341.

32. Ashton, N.J., et al., A multicentre validation study of the diagnostic value of plasma neurofilament light. Nat Commun, 2021. 12(1): p. 3400.

33. Chelban, V., et al., Neurofilament light levels predict clinical progression and death in multiple system atrophy. Brain, 2022. 145(12): p. 4398–4408.

34. Lisman, J., et al., Viewpoints: how the hippocampus contributes to memory, navigation and cognition. Nat Neurosci, 2017. 20(11): p. 1434–1447.

35. Escartin, C., et al., Reactive astrocyte nomenclature, definitions, and future directions. Nat Neurosci, 2021. 24(3): p. 312–325.

36. Ouyang, X., et al., Peripheral biomarkers of neuroinflammation in Alzheimer’s disease and vascular dementia. Biochem Pharmacol, 2026. 250(Pt 2): p. 118077.

37. Ossenkoppele, R., et al., Amyloid and tau PET-positive cognitively unimpaired individuals are at high risk for future cognitive decline. Nat Med, 2022. 28(11): p. 2381– 2387.

38. Horie, K., et al., Plasma MTBR-tau243 biomarker identifies tau tangle pathology in Alzheimer’s disease. Nature Medicine, 2025. 31(6): p. 2044–2053.

39. Stern, Y., et al., A framework for concepts of reserve and resilience in aging. Neurobiol Aging, 2023. 124: p. 100–103.

40. Svenningsson, A.L., et al., Biological mechanisms of resilience to tau pathology in Alzheimer’s disease. Alzheimers Res Ther, 2024. 16(1): p. 221.

41. de Vries, L.E., et al., The concept of resilience to Alzheimer’s Disease: current definitions and cellular and molecular mechanisms. Mol Neurodegener, 2024. 19(1): p. 33.

42. O’Bryant, S.E., et al., Validation of the new interpretive guidelines for the clinical dementia rating scale sum of boxes score in the national Alzheimer’s coordinating center database. Arch Neurol, 2010. 67(6): p. 746–9.

43. Chong, J.R., et al., *Plasma P-tau181 to A*β*42 ratio is associated with brain amyloid burden and hippocampal atrophy in an Asian cohort of Alzheimer’s disease patients with concomitant cerebrovascular disease*. Alzheimers Dement, 2021. 17(10): p. 1649–1662.

44. Saridin, F.N., et al., Cerebrovascular disease in suspected non-Alzheimer’s pathophysiology and cognitive decline over time. Eur J Neurol, 2022. 29(7): p. 1922– 1929.

