## Supplemental for "Integrating Plasma Brain-Derived pTau217, White Matter Hyperintensity and Hippocampal Volumes as Optimized Prognostic Markers for Cognitive Decline in an Asian Cohort with High Cerebrovascular Disease Burden"

### Supplementary Materials

#### Supplemental Figure 1. Flow Chart of Study Participants n = 523

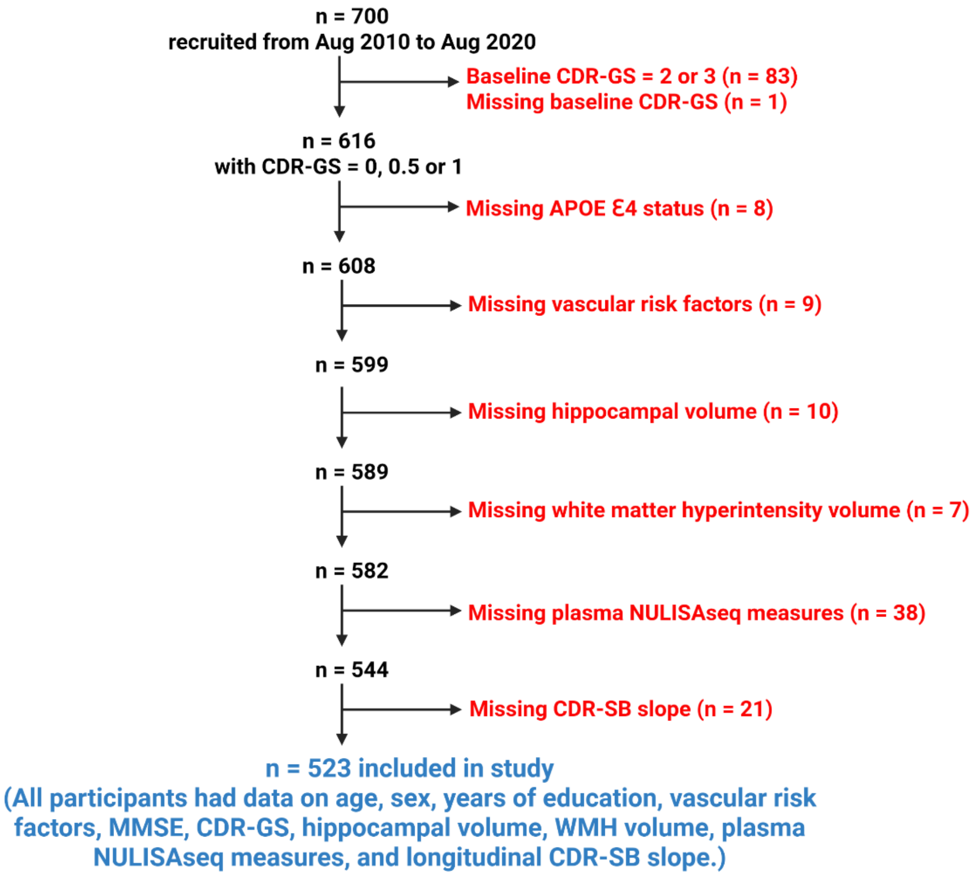

**Supplemental Figure 2. Longitudinal Trajectories of Clinical Dementia Rating Sum of Boxes According to Number of Positive AVN Biomarkers (Based on Singleplex BD-pTau217, Fazekas score, and Medial Temporal Atrophy Score)**

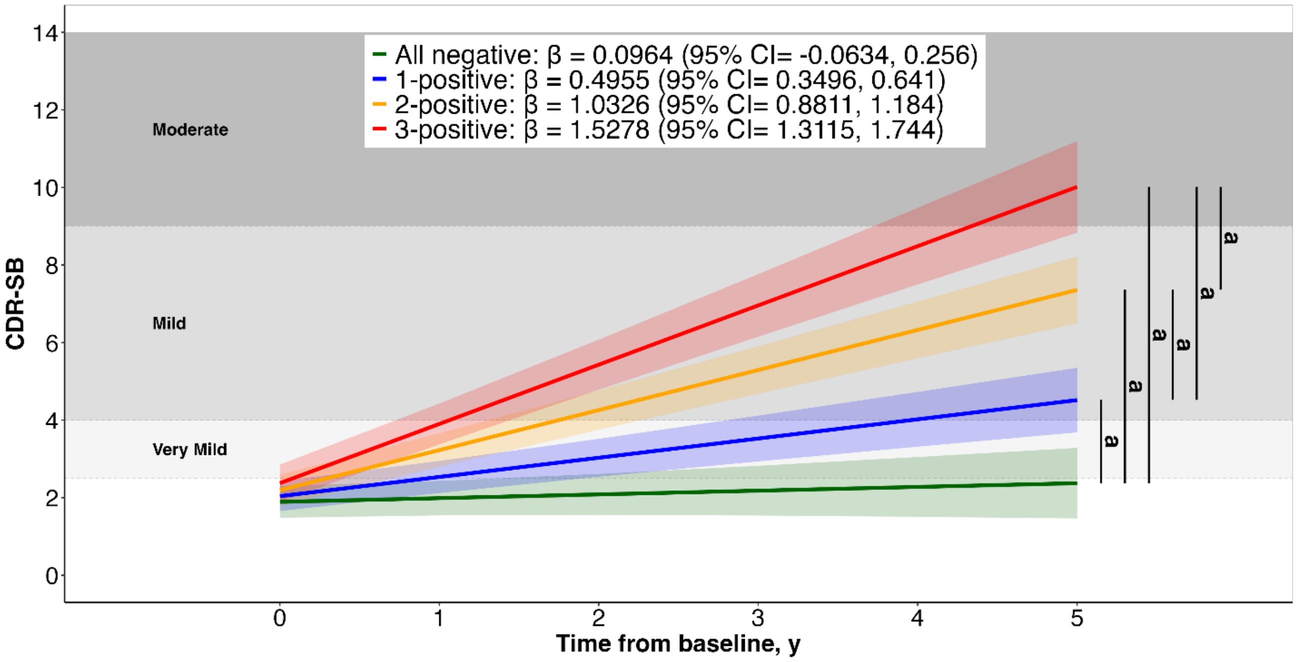


Supplemental Figure 2. Trajectory plots indicate the mean longitudinal trajectories (solid line) of clinical dementia rating sum of boxes (CDR-SB), and associated 95% confidence intervals (shading), estimated with linear mixed-effects model. Trajectories are stratified based on the number of positive AVN biomarkers (All negative [green], 1-positive [blue], 2-positive [orange], 3-positive [red]), modelled with an interaction between group and time. Participants were stratified based on singleplex BD-pTau217 (A), Fazekas score (V), and medial temporal atrophy score (N). Participants who were negative for all three biomarkers (A-V-N-) were classified as "All negative". Participants who were positive for one biomarker (A+V-N-, A-V+N-, or A-V-N+) were collectively classified as "1-positive". Participants who were positive for two biomarkers (A+V+N-, A+V-N+, or A-V+N+) were collectively classified as "2-positive". Participants who were positive for all three biomarkers (A+V+N+) were classified as "3-positive". Refer to Supplemental Table 1 for the threshold used to define positivity for each biomarker. Model included random slopes and intercepts and was adjusted for age, sex, years of education, *APOE4* status, hypertension, hyperlipidemia, diabetes, and cardiovascular disease status, and baseline clinical dementia rating global score (CDR-GS). Shaded background regions indicate dementia severity categories based on CDR-SB thresholds[1]: 2.5–4.0 (very mild), 4.5–9.0 (mild), 9.5–15.5 (moderate), and 16.0–18.0 (severe). *P* values represent post hoc pairwise comparing the slopes between groups.
[a] *P* < 0.001

### Supplemental Figure 3. Longitudinal Trajectories of Clinical Dementia Rating Sum of Boxes According to AVN Biomarkers

#### (A) Based on Multiplex BD-pTau217, White Matter Hyperintensity and Hippocampal volumes (n = 523)

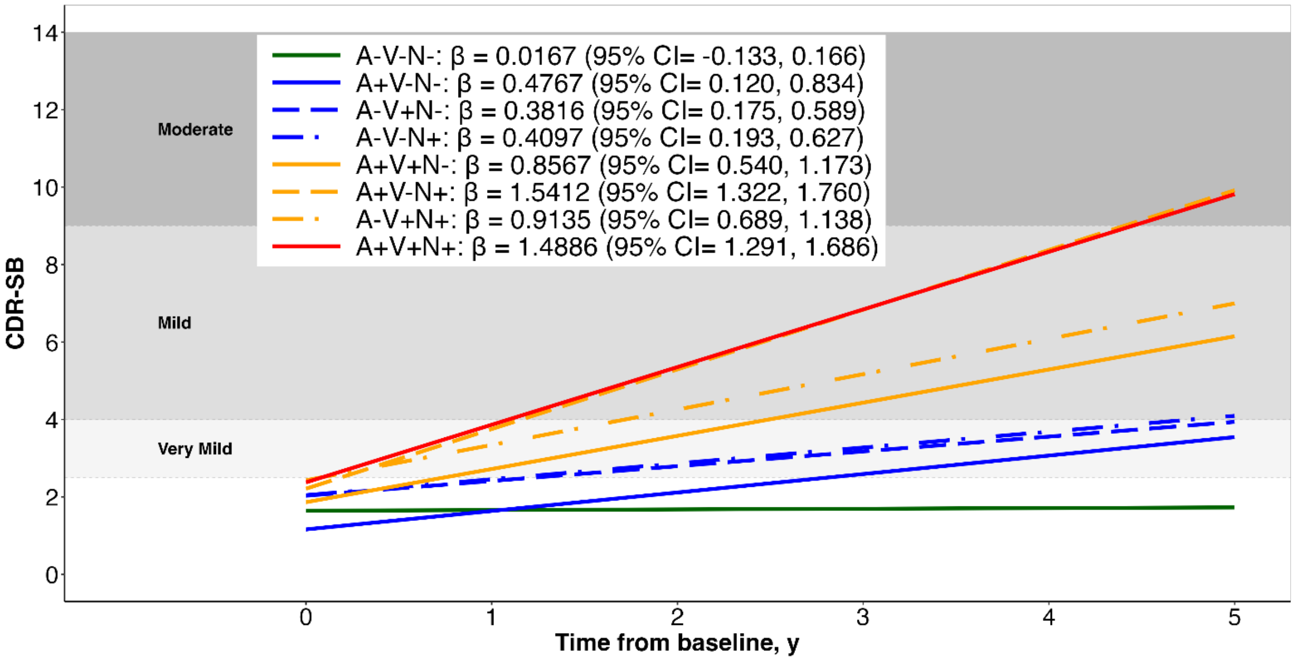


#### (B) Based on Singleplex BD-pTau217, Fazekas and Medial Temporal Atrophy scores (n = 519)

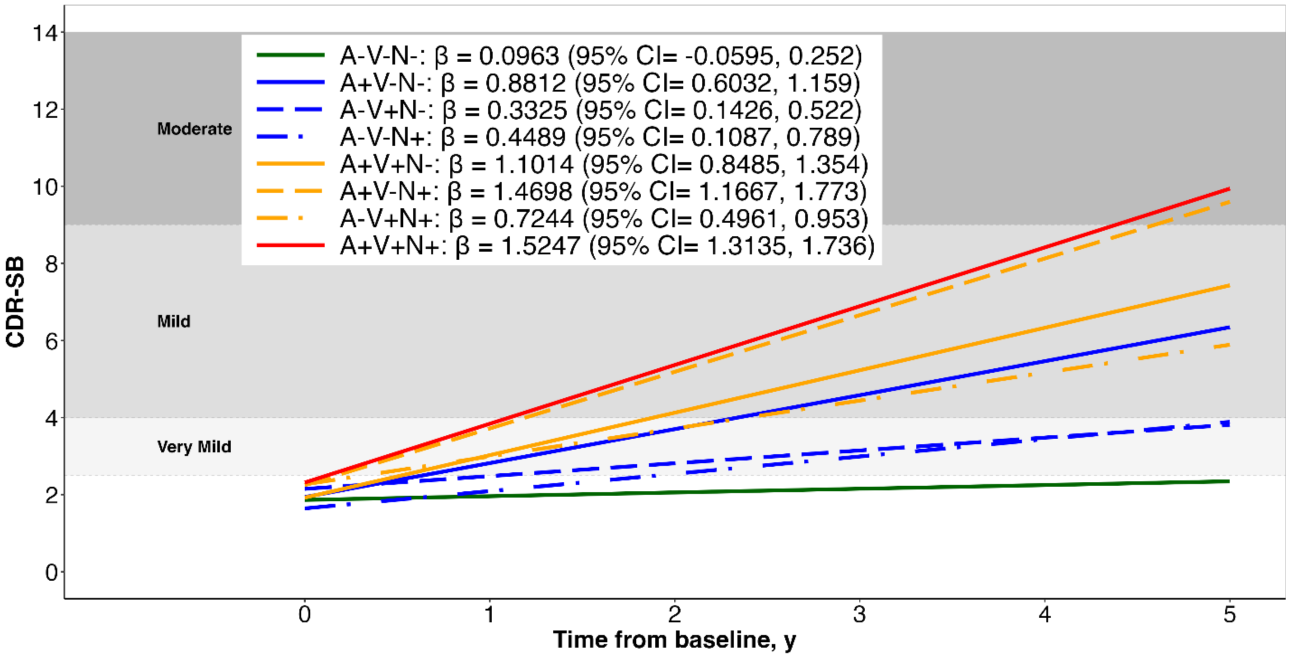


Supplemental Figure 3. Trajectory plots indicate the mean longitudinal trajectories of clinical dementia rating sum of boxes (CDR-SB), and associated 95% confidence intervals, estimated with linear mixed-effects models. Trajectories are stratified based on the eight AVN combinations (A-V-N-, A+V-N-, A-V+N-, A-V-N+, A+V+N-, A+V-N+, A-V+N+, A+V+N+), modelled with an interaction between group and time. Models included random slopes and intercepts and were adjusted for age, sex, years of education, *APOE4* status, hypertension, hyperlipidemia, diabetes, and cardiovascular disease status, and baseline clinical dementia rating global score (CDR-GS). Shaded background regions indicate dementia severity categories based on CDR-SB thresholds [1]: 2.5–4.0 (very mild), 4.5–9.0 (mild), 9.5–15.5 (moderate), and 16.0–18.0 (severe).

### Supplemental Table 1. Dichotomization of Plasma BD-pTau217, White Matter Hyperintensity and Hippocampal volumes

**(A) Multiplex plasma BD-pTau217 (n = 213)**

| Risk groups | Specificity (%) | Sensitivity (%) | Participants in each risk group (n (%)) | Aβ PET negative (A-; n (%)) | Aβ PET positive (A+; n (%)) |
|---|---|---|---|---|---|
| **Plasma BD-pTau217 (NPQ)** | | | | | |
| **Low-risk (<11.691)** | 92.1 | 91.9 | 134 (63) | 128 (96) | 6 (4) |
| **High-risk (>=11.691)** | | | 79 (37) | 11 (14) | 68 (86) |

**(B) Singleplex plasma BD-pTau217 (n = 213)**

| Risk groups | Specificity (%) | Sensitivity (%) | Participants in each risk group (n (%)) | Aβ PET negative (A-; n (%)) | Aβ PET positive (A+; n (%)) |
|---|---|---|---|---|---|
| **Plasma BD-pTau217 (pg/mL)** | | | | | |
| **Low-risk (<0.411)** | 90.6 | 91.9 | 132 (62) | 126 (95) | 6 (5) |
| **High-risk (>=0.411)** | | | 81 (38) | 13 (16) | 68 (84) |

**(C) WMH volumes (n = 669)**

| Risk groups | Specificity (%) | Sensitivity (%) | Participants in each risk group (n (%)) | Fazekas score <2 (V-; n (%)) | Fazekas score ≥2 (V+; n (%)) |
|---|---|---|---|---|---|
| **WMH volume (%)** | | | | | |
| **Low-risk (<0.3726)** | 92.3 | 80.6 | 328 (49) | 251 (77) | 77 (23) |
| **High-risk (>=0.3726)** | | | 341 (51) | 21 (6) | 320 (94) |

**(D) Hippocampal volumes (n = 676)**

| Risk groups | Specificity (%) | Sensitivity (%) | Participants in each risk group (n (%)) | MTA score <2 (N-; n (%)) | MTA score ≥2 (N+; n (%)) |
|---|---|---|---|---|---|
| **Hippocampal volume (%)** | | | | | |
| **Low-risk (>0.4859)** | 66.9 | 81.6 | 297 (44) | 238 (80) | 59 (20) |
| **High-risk (<=0.4859)** | | | 379 (56) | 118 (31) | 261 (69) |

Supplemental Table 1. Dichotomization of multiplex plasma BD-pTau217 (A), singleplex plasma BD-pTau217 (B), white matter hyperintensity (WMH) volumes (C), and hippocampal volumes (D). Thresholds are indicated in parenthesis next to the low- and high-risk groups. For each biomarker, the second and third column correspond to the specificity and sensitivity respectively, while the fourth column corresponds to the number of individuals falling in each risk category. A/V/N status is shown for the low- and high-risk groups. The percentage of A-/V-/N- in the low-risk group and the percentage of A+/V+/N+ in the high-risk group correspond to each evaluated threshold's negative predictive value (NPV) and positive predictive value (PPV), respectively.

(A) and (B): To determine the binary reference of multiplex plasma BD-pTau217 (A) or singleplex plasma BD-pTau217 (B) for Aβ positron emission tomography (PET) positivity (A+), a subset of the main cohort with available measurements for both plasma biomarkers and Aβ PET was used (n = 213). Details of the procedures for measuring brain amyloid burden have been published previously [2, 3]. Briefly, imaging was performed using [$^{11}$C]Pittsburgh Compound B (PiB, n = 184) or [$^{18}$F]flutafuranol (n = 29) amyloid tracer radioligands. Each [$^{11}$C]-PiB or [$^{18}$F]-flutafuranol image was independently visually interpreted by three experienced raters blinded to the clinical diagnosis of each participant, and was used to classify individuals as Aβ positive (A+) or Aβ negative (A-). For multiplex plasma BD-pTau217, threshold was determined by maximizing the Youden index and published [2]. For singleplex plasma BD-pTau217, threshold was derived by setting at 90% specificity while maximizing sensitivity.

(C): To determine the binary reference of WMH volumes for significant WMH burden (V+), a subset of the main cohort with available WMH volume measurements and Fazekas scores was used (n = 669). Briefly, WMH were visually graded, and defined as hyperintense on FLAIR sequences and hypointense on T1-weighted images, using the modified Fazekas scale focusing on deep white matter[4]. Significant WMH burden (V+) was defined as Fazekas score ≥2 [4]. Threshold was determined by maximizing the Youden index.

(D): To determine the binary reference of hippocampal volumes for significant neurodegeneration (N+), a subset of the main cohort with available hippocampal volume measurements and medial temporal atrophy (MTA) scores was used (n = 676). Briefly, MTA was graded on coronal T1-weighted slices, according to the Scheltens scale [4]. Significant neurodegeneration (N+) was defined as MTA score ≥2 [4]. Threshold was determined by maximizing the Youden index.

### Supplemental Table 2. Stratification of Participants into AVN Groupings

| AVN Group | Number of positive biomarkers | Based on multiplex BD-pTau217(A), white matter hyperintensity (V) and hippocampal (N) volumes (n = 523) | Based on singleplex BD-pTau217(A), Fazekas (V) and medial temporal atrophy (N) scores (n = 519) |
|---|---|---|---|
| **A-V-N-** | **All negative** | 128 | 126 |
| **A+V-N-** | **1-positive** | 22 | 40 |
| **A-V+N-** | | 70 | 88 |
| **A-V-N+** | | 62 | 30 |
| **A+V+N-** | **2-positive** | 31 | 52 |
| **A+V-N+** | | 63 | 36 |
| **A-V+N+** | | 64 | 67 |
| **A+V+N+** | **3-positive** | 83 | 80 |

Supplemental Table 2. The table shows the AVN combinations and corresponding participant numbers, stratified based on multiplex BD-pTau217 (A), white matter hyperintensity volume (V), and hippocampal volume (N) (third column; n = 523), or singleplex BD-pTau217 (A), Fazekas score (V), and medial temporal atrophy score (N) (fourth column; n = 519). In the second column, participants who were negative for all three biomarkers (A-V-N-) were classified as "All-negative". Participants who were positive for one biomarker (A+V-N-, A-V+N-, or A-V-N+) were collectively classified as "1-positive". Participants who were positive for two biomarkers (A+V+N-, A+V-N+, or A-V+N+) were collectively classified as "2-positive". Participants who were positive for all three biomarkers (A+V+N+) were classified as "3-positive". Refer to Supplemental Table 1 for the threshold used to define positivity for each biomarker.

**Supplemental Table 3. Cox Regression Analyses of Risk for Incident Cognitive Decline in Participants Stratified by Number of Positive AVN Biomarkers (Based on Singleplex BD-pTau217, Fazekas score, and Medial Temporal Atrophy Score)**

| Incident Cognitive Decline (n = 519) | | | | | |
|---|---|---|---|---|---|
| | **Stable, n (%)** | **Declined, n (%)** | **Hazard ratio** | **95% CI** | **p-value** |
| **All negative (n = 126):**<br>A-V-N- (n = 126) | 104 (82) | 22 (18) | 1.00 (Reference) | | |
| **1-positive (n = 158):**<br>A+V-N- (n = 40)<br>A-V+N- (n = 88)<br>A-V-N+ (n = 30) | 96 (61) | 62 (39) | 2.482 | 1.501, 4.104 | <0.001 |
| **2-positive (n = 155):**<br>A+V+N- (n = 52)<br>A+V-N+ (n = 36)<br>A-V+N+ (n = 67) | 66 (43) | 89 (57) | 3.881 | 2.319, 6.494 | <0.001 |
| **3-positive (n = 80):**<br>A+V+N+ (n = 80) | 29 (36) | 51 (64) | 4.741 | 2.656, 8.465 | <0.001 |

Supplemental Table 3. Cox regression with hazard ratios and 95% confidence intervals. Incident cognitive decline was defined as an increase in CDR-GS of ≥0.5 at annual follow-up compared with the baseline. Participants were stratified by the number of positive AVN biomarkers, based on singleplex BD-pTau217 (A), Fazekas score (V), and medial temporal atrophy score (N). Participants who were negative for all three biomarkers (A-V-N-) were classified as "All negative". Participants who were positive for one biomarker (A+V-N-, A-V+N-, or A-V-N+) were collectively classified as "1-positive". Participants who were positive for two biomarkers (A+V+N-, A+V-N+, or A-V+N+) were collectively classified as "2-positive". Participants who were positive for all three biomarkers (A+V+N+) were classified as "3-positive". Refer to Supplemental Table 1 for the threshold used to define positivity for each biomarker. Cox regression model was adjusted for age, sex, years of education, *APOE4* status, hypertension, hyperlipidemia, diabetes, and cardiovascular disease status, and baseline clinical dementia rating global score (CDR-GS).

### Supplemental Table 4. Cox Regression Analyses of Risk for Incident Cognitive Decline in Participants Stratified by AVN Biomarkers

#### (A) Based on Multiplex BD-pTau217, White Matter Hyperintensity and Hippocampal volumes

| | Incident Cognitive Decline (n = 523) | | | | |
|---|---|---|---|---|---|
| | **Stable, n (%)** | **Declined, n (%)** | **Hazard ratio** | **95% CI** | **p-value** |
| **A-V-N- (n = 128)** | 110 (86) | 18 (14) | 1.00 (Reference) | | |
| **A+V-N- (n = 22)** | 14 (64) | 8 (36) | 2.357 | 1.015, 5.475 | 0.046 |
| **A-V+N- (n = 70)** | 42 (60) | 28 (40) | 3.300 | 1.800, 6.050 | <0.001 |
| **A-V-N+ (n = 62)** | 43 (69) | 19 (31) | 2.524 | 1.288, 4.945 | 0.007 |
| **A+V+N- (n = 31)** | 13 (42) | 18 (58) | 5.366 | 2.681, 10.738 | <0.001 |
| **A+V-N+ (n = 63)** | 13 (21) | 50 (79) | 9.140 | 5.019, 16.647 | <0.001 |
| **A-V+N+ (n = 64)** | 36 (56) | 28 (44) | 4.113 | 2.135, 7.920 | <0.001 |
| **A+V+N+ (n = 83)** | 26 (31) | 57 (69) | 7.122 | 3.884, 13.061 | <0.001 |

#### (B) Based on Singleplex BD-pTau217, Fazekas and Medial Temporal Atrophy scores (n = 519)

| | Incident Cognitive Decline (n = 519) | | | | |
|---|---|---|---|---|---|
| | **Stable, n (%)** | **Declined, n (%)** | **Hazard ratio** | **95% CI** | **p-value** |
| **A-V-N- (n = 126)** | 104 (82) | 22 (18) | 1.00 (Reference) | | |
| **A+V-N- (n = 40)** | 19 (47) | 21 (53) | 3.228 | 1.743, 5.975 | <0.001 |
| **A-V+N- (n = 88)** | 56 (64) | 32 (36) | 2.261 | 1.296, 3.944 | 0.004 |
| **A-V-N+ (n = 30)** | 21 (70) | 9 (30) | 1.838 | 0.814, 4.151 | 0.143 |
| **A+V+N- (n = 52)** | 17 (33) | 35 (67) | 4.293 | 2.409, 7.651 | <0.001 |
| **A+V-N+ (n = 36)** | 6 (17) | 30 (83) | 6.296 | 3.404, 11.647 | <0.001 |
| **A-V+N+ (n = 67)** | 43 (64) | 24 (36) | 2.400 | 1.279, 4.504 | 0.006 |
| **A+V+N+ (n = 80)** | 29 (36) | 51 (64) | 4.680 | 2.610, 8.391 | <0.001 |

Supplemental Table 4. Cox regression with hazard ratios and 95% confidence intervals. Incident cognitive decline was defined as an increase in CDR-GS of ≥0.5 at annual follow-up compared with the baseline. Cox regression model was adjusted for age, sex, years of education, *APOE4* status, hypertension, hyperlipidemia, diabetes, and cardiovascular disease status, and baseline clinical dementia rating global score (CDR-GS).

(A): Participants were stratified based on multiplex BD-pTau217 (A), white matter hyperintensity volume (V), and hippocampal volume (N). Refer to eTable 1 for the threshold used to define positivity for each biomarker.

(B): Participants were stratified based on singleplex BD-pTau217 (A), Fazekas score (V), and medial temporal atrophy score (N). Refer to Supplemental Table 1 for the threshold used to define positivity for each biomarker.

### References


1. O'Bryant, S.E., et al., *Validation of the new interpretive guidelines for the clinical dementia rating scale sum of boxes score in the national Alzheimer's coordinating center database.* Arch Neurol, 2010. **67**(6): p. 746–9.
2. Chong, J.R., et al., *Plasma brain-derived p-Tau217 outperforms other p-Tau species in detecting abnormal brain amyloid in an Asian cohort of older people with cerebrovascular disease burden.* The Journal of Prevention of Alzheimer's Disease, 2026. **13**(7): p. 100615.
3. Chong, J.R., et al., *Plasma P-tau181 to Aβ42 ratio is associated with brain amyloid burden and hippocampal atrophy in an Asian cohort of Alzheimer's disease patients with concomitant cerebrovascular disease.* Alzheimers Dement, 2021. **17**(10): p. 1649–1662.
4. Saridin, F.N., et al., *Cerebrovascular disease in suspected non-Alzheimer's pathophysiology and cognitive decline over time.* Eur J Neurol, 2022. **29**(7): p. 1922–1929.
